# Molecular surveillance detects high prevalence of the neglected parasite *Mansonella ozzardi* in the Colombian Amazon

**DOI:** 10.1101/2023.05.10.23289806

**Authors:** KJ Dahmer, M Palma-Cuero, K Ciuoderis, C Patiño, S Roitman, Z Li, A Sinha, JL Hite, O Bellido Cuellar, JP Hernandez-Ortiz, JE Osorio, BM Christensen, CKS Carlow, M Zamanian

## Abstract

Mansonellosis is an undermapped insect-transmitted disease caused by filarial nematodes that are estimated to infect hundreds of millions of people globally. Despite their prevalence, there are many outstanding questions regarding the general biology and health impacts of the responsible parasites. Historical reports suggest that the Colombian Amazon is endemic for mansonellosis and may serve as an ideal location to pursue these questions in the backdrop of other endemic and emerging pathogens. We deployed molecular and classical diagnostic approaches to survey *Mansonella* prevalence among adults belonging to indigenous communities along the Amazon River and its tributaries near Leticia, Colombia. Deployment of a loop-mediated isothermal amplification (LAMP) assay on blood samples revealed an infection prevalence of ∼40% for *Mansonella ozzardi*. This assay identified significantly more infections than blood smear microscopy or LAMP assays performed using plasma, likely reflecting greater sensitivity and the ability to detect low microfilaremias or occult infections. *Mansonella* infection rates increased with age and were higher among males compared to females. Genomic analysis confirmed the presence of *M. ozzardi* that clusters closely with strains sequenced in neighboring countries. We successfully cryopreserved and revitalized *M. ozzardi* microfilariae, advancing the prospects of rearing infective larvae in controlled settings. These data suggest an underestimation of true mansonellosis prevalence, and we expect that these methods will help facilitate the study of mansonellosis in endemic and laboratory settings.

## Introduction

Mansonellosis is a highly prevalent but undermapped [1] and understudied parasitic disease that infects hundreds of millions of people throughout Africa and Central and South America [2]. Three species of insect-transmitted parasitic nematodes (*Mansonella ozzardi, M. perstans* and *M. streptocerca*) are responsible for the majority of human cases although other *Mansonella* species have the potential to infect humans, including the recently discovered *Mansonella* sp. “DEUX” which has revitalized demands for allocating resources to study this severely neglected disease [2–5].

There are major gaps in our understanding of the basic biology and clinical or subclinical impacts of mansonellosis in human populations. *Mansonella* infections underlie variable clinical presentations that are likely underrecognized [4] and may alter host immunity in ways that alter vaccine responses and susceptibility to other pathogens [6–8]. Drugs used in mass drug administration campaigns for lymphatic filariasis and onchocerciasis are not as effective against *Mansonella spp*. [9–13] suggesting a unique genetic basis for anthelmintic resistance [14–16]. Additionally, *Mansonella* infections can introduce diagnostic challenges related to species mis-identification [17,18], cross-reactivity of immunochromatographic tests [19,20], and the inability to easily discern occult or amicrofilaremic infections [21]. Together, these limitations can potentially confound parasite elimination and surveillance programs focused on more prominent filarial worm parasites in co-endemic regions [9–13,16].

To address these challenges, we deployed both classical and molecular diagnostic approaches to carry out more precise epidemiological surveys of *Mansonella* prevalence in adults from villages along the Amazon River and its tributaries around the capital city of Leticia in the Amazonas Department of Colombia. The Amazon basin is historically associated with “new world” *M. ozzardi* [22–25] and is ideally situated to pursue fundamental questions about *Mansonella* biology, clinical presentations, and interactions with other pathogens that are endemic in the region. Parasite surveys in this region are outdated [22–24], but incidental findings from community health workers in the course of malaria screening and estimates from communities in neighboring countries [4,21,26] suggest a high prevalence.

The gold standard diagnostic for *M. perstans* and *M. ozzardi* is blood smear microscopy, which requires morphological differentiation of the circulating microfilariae (mf) stage [4,17]. Molecular diagnostic approaches including real-time PCR (RT-PCR) [27–29] are largely restricted to laboratory settings. However, more recently developed loop-mediated isothermal amplification (LAMP) assays with a simple colorimetric readout are more amenable to field studies [30]. We comparatively profiled LAMP assays with classical microscopy-based techniques to evaluate potential underestimation of parasite prevalence among a cluster of indigenous communities in the Colombian Amazon region. Microfilariae were isolated for population genomic analyses of *Mansonella* with respect to previously characterized field isolates, as well as for the establishment of a cryopreservation protocol to enable laboratory studies. Lastly, we probed associations of *Mansonella* infection status with other clinical and demographic variables. We expect that these data and methods will help facilitate future studies of *Mansonella*.

## Methods

### Study design and sampling procedure

This study was conducted between 2021 and 2023 in the Amazonas Department of Colombia in areas surrounding the capital city of Leticia and Puerto Nariño municipalities (**Figure 1A**). This region of vast biodiversity is largely populated by indigenous communities situated near the Amazon River and its tributaries. The study protocol was reviewed and approved by the UW-Madison IRB (study # 2019-1107) and Corporación para Investigaciones Biológicas (CIB #17022021). The study population (n = 235) from 13 communities was recruited by convenience and individuals ≥18 years of age were invited to participate and enrolled after providing written informed consent. Peripheral (venous) blood samples were collected into ethylenediaminetetraacetic acid (EDTA) tubes and thin and thick blood smears were prepared and examined for the presence of filarial parasites and malaria. Serum/plasma were extracted and aliquoted along with whole blood into 1.5 mL tubes and stored at -80°C for detection of *Mansonella* and other pathogens (**Figure 1B**). Self-reported socio-demographic and epidemiological data were collected using a structured questionnaire through face-to-face interviews at the enrollment site. Recorded data included demographics (age, sex, ethnicity, occupation, place of residence), housing conditions, travel history, medical history (pre-existing diseases), and clinical symptoms.

**Figure 1.**
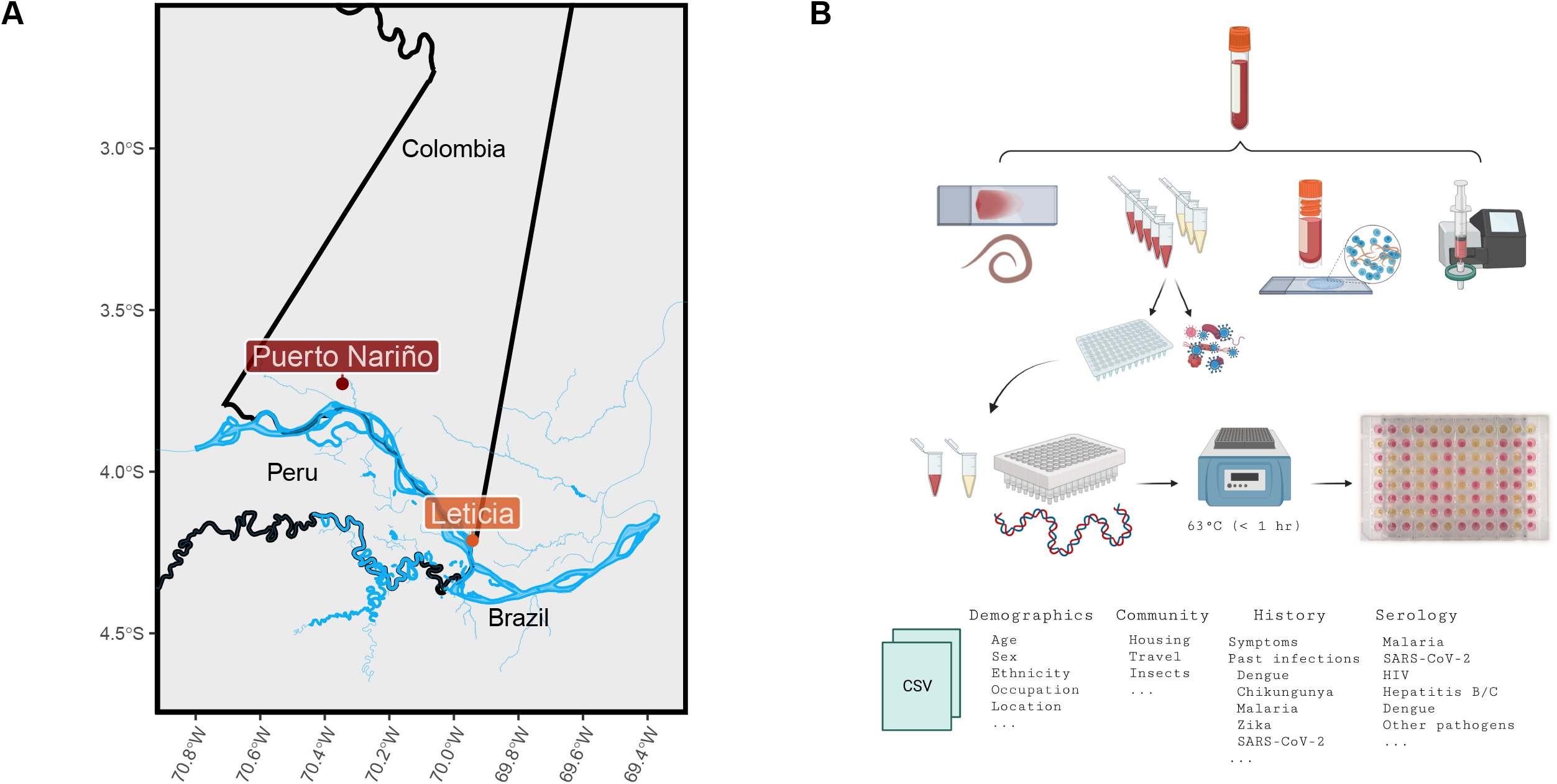
**A)** Map of the study region in the Amazonas Department of Colombia depicting the capital city of Leticia and the Puerto Nariño municipality where samples were collected across three surveys. The study region is adjacent to the Amazon River at the borders of Colombia, Brazil, and Peru. **B)** Schematic of sampling efforts and endpoints for blood samples acquired from adult volunteers. Blood smears and preparations of cryopreserved samples were conducted in the field. Whole blood was obtained in EDTA tubes and transferred on ice to a laboratory in Leticia. Aliquots of whole-blood and plasma were prepared from samples and used for downstream serology, molecular diagnostics, and genomic analyses. Questionnaires were filled out at the time of sampling.

### Microscopy smears and microfilariae quantification

Routine thin and thick smears were prepared at the time of blood draws. Slides were allowed to dry at room temperature, Giemsa stained and examined microscopically. For quantitative microscopy, 40 μL of thawed blood was mixed with equal amounts of water and microfilaria (mf) were counted using light microscopy. For filtration of thawed blood aliquots, 1 mL of blood mixed with 4 ml of phosphate-buffered saline (PBS) was added to a Millipore syringe (Sigma-Aldrich, Z268429) with a 5 μm filter membrane (Millipore Sigma, TMTP02500). Filter membranes were imprinted in a microscope slide, allowed to dry before methanol fixation and Giemsa staining for microscopical observation.

### Cryopreservation and recovery of microfilariae

Blood samples were transported on ice from Puerto Nariño to Leticia, where they were mixed with 5% dimethyl sulfoxide (DMSO) at a 1:1 ratio, aliquoted in cryogenic tubes and stored at - 80°C [31]. Processed samples were then shipped to Medellin and stored at -80°C. Samples were thawed in a water bath at 37°C for 5 minutes and washed twice with sterile PBS (at 37°C), followed by centrifugation at 2,000 rpm for 10 minutes. The mf pellet was gently mixed with 2 mL of RPMI-1640 + 1x Antibiotic-Antimycotic (Gibco, 15240096). 1 mL aliquots were added to wells in a 6-well cell culture plate and the plate was incubated at 37°C and 5% CO_2_. Movement patterns of mf were recorded using an inverted microscope (40X objective). Movement levels were quantified using a customized optical flow pipeline[32].

### DNA extraction from blood and plasma samples

DNA was extracted from whole blood and plasma using the Quick-DNA 96-plus kit (Zymo, D4070/71) with minor adjustments to the protocol. For blood samples, the “solid tissue protocol” was followed by adding 50 μL of blood in place of water and tissue. After incubation for 2.5 hours, samples were spun at 3,500g for 2 minutes and supernatant lysate was transferred to the Zymo-spin 96-XL plate. For plasma samples, the “biological fluid and cells” protocol was followed. Final DNA was eluted with 15 μL of water and concentrations were assessed via NanoDrop for quality assurance.

### Loop-mediated isothermal amplification assays

LAMP reactions were carried out as previously described [30] with slight modifications. Briefly, each LAMP reaction contained 1.6 μM each of primers FIP (5’-CGCAAACAGAAGCCCGAAAC-GCTCGCAATTTCATAGTGG-3’) and BIP (5’-CTTGCGCGTAGCATTAGATCC-TCCGAAATGTATACGACAGAT-3’), 0.2 μM each of F3 (5’-GCACGAAATGTTTTTGTACG-3’) and B3 (5’-CGTATCACCGTTGATGACG-3’), 0.4 μM each of LF (5’-AAGCCTAAGCCTAAGCCTGA-3’) and LB (5’-GCACATCTTCAATCTCCTCTTGC-3’), 2 μl 10X GuHCL, 10 μl of WarmStart Colorimetric LAMP 2x Master Mix (NEB, M1804L), 4 μl water and 2 μl of template DNA, or 2 μl water for non-template controls for a total volume of 20 μl per reaction. For a colorimetric readout, reactions were incubated at 63°C for 30 minutes in a SimpliAmp Thermocycler (Applied Biosystems, A24811) and images were acquired using an ImageQuant 800 (Cytiva). A post amplification color of yellow indicated detection of *M. ozzardi*, and pink (or orange) indicated no detection.

For semi-quantitative LAMP (sq-LAMP), simultaneous colorimetric and fluorescent readouts were obtained by adding SYTO™ 9 green (Invitrogen, S34854) to a final concentration of 1 μM in the colorimetric LAMP reaction. Reactions were performed in a Bio-Rad CFX Opus 96 Real-Time PCR instrument at 65°C with total fluorescence read in the SYBR/FAM channel every 15 seconds for 150 “cycles” (∼53 minutes with each “cycle” corresponding to 21.2 seconds of reaction time, 15 seconds combined with plate reading time). A cut-off time threshold (Tt) value of 30 minutes was used to differentiate positive/negative reactions, with Tt defined as the time (min) to reach the fluorescence detection threshold. A Tt ≤ 30 minutes indicated the detection of the target, whereas a Tt > 30 minutes or N/A, indicated no detection. Plates were scanned using the Epson Perfection v600 Photo Scanner.

### DNA extraction and Illumina library construction and sequencing

DNA was extracted from 200 uL of whole blood using the MagAttract HMW DNA kit (Qiagen, 67563) following the manufacturer’s instructions. The NEBNext Microbiome DNA enrichment kit (NEB, E2612) was used as directed to enrich *Mansonella* DNA and reduce human DNA prior to library construction. The Illumina libraries were constructed using the NEBNext Ultra II DNA Library Prep Kit for Illumina (NEB, E7645) as described by the manufacturer. The quality and concentration of each library were determined using a 2100 Bioanalyzer with a high-sensitivity DNA chip (Agilent Technologies). Libraries were diluted to 1□nM with 10□mM Tris, 0.1□mM EDTA pH 8. Phi X DNA (5%) was added to balance base pair composition in these A:T rich filarial libraries prior to sequencing on a NovaSeq platform (paired end, 150□bps).

### Bioinformatic analysis

Raw Illumina reads were processed to remove adapters and poor-quality reads using the BBTools package (https://jgi.doe.gov/data-and-tools/bbtools/). For each isolate, the reads were mapped to a combined reference sequence set comprising a *M. ozzardi* reference genome from Brazil [16], a reference mitogenome KX822021.1 [33] and the *Wolbachia w*Moz assembly GCF_020278625.1 [34] using bowtie2 [35]. Reads mapping to mitochondria were extracted from the bam alignment files using samtools [36] and assembled into circular mitochondrial genomes using GetOrganelle v1.7.7.0 [37]. The average nucleotide identity scores between pairs of all assembled mitogenomes and published mitogenomes, namely the accessions KX822021.1 and MN416134.1 for *M. ozzardi* [21,33] and MT361687.1 and MN432521.1 for *M. perstans* [21,38] were calculated using the OrthoANIu tool [39]. Multiple sequence alignments of all *M. ozzardi* and *M. perstans* mitogenomes were obtained using mafft v7.149b [40]. The phylogenetic tree based on this alignment was generated using the iqtree online server [41], which performs automatic best fit substitution model selection using ModelFinder [42]. Bootstrap support values were calculated based on ultrafast bootstrap [43] with 1000 replicates. The tree was annotated on the iTOL webserver [44].

### Serological tests for viral infections

Serum samples were used for antibody detection against several pathogens. Anti-dengue immunoglobulin G (IgG) and IgM were detected using SD BIOLINE Dengue Duo rapid test (Abbott), following manufacturer’s instructions. Anti-Human Immunodeficiency Virus (HIV), Hepatitis B Virus (HBV), and Hepatitis C Virus (HCV) as well as anti-IgM against Severe Acute Respiratory Syndrome Coronavirus 2 (SARS-COV-2) were detected by chemiluminescence immunoassay using the Architect I1000 system (Abbott), following manufacturer’s instructions.

### Data analysis

Self-reported data were recorded on printed surveys by study personnel and completed forms were entered into Microsoft Excel by double entry. Excel spreadsheets were used for data inspection, cleaning, and quality control before data analysis using R Studio software v4.2.2 (42). To examine sex- and age-based differences of *M. ozzardi* infections, we used a linear model (lm) to get the residuals and assess the normality of our data. We determined our data did not follow a normal distribution (Shapiro-Wilk test, male age: W = 0.93224, p-value = 0.0002649, female age: W = 0.89475, p-value = 8.05e-09). Based on these results, we ran a non-parametric Wilcox rank sum test with a p-value < 0.05 considered statistically significant. Next, to examine differences between infection prevalence across demographic variables and analyses on reported symptoms and serology results, we used generalized linear models (GLMs) with binomial distributions and log link functions [45]. We conducted model selection analyses using the *aictab* function in the R package AICcmodavg [46]. We built candidate models starting with the full model with all combinations of main effects among relevant biological and methodological factors, while avoiding overfitting. We compared candidate models using Akaike’s information criterion and ΔAIC (the difference in AIC values for the focal model and the model with the lowest AIC, i.e., the ‘winning’ model) [47]. We also calculated the Akaike weight (𝓌), which further quantifies the probability that a model is the most appropriate model relative to the candidate models (**Supplementary Table 1**). ΔAIC less than two and a higher 𝓌 generally indicates that a model has substantial support while a suite of best models with low weights (𝓌 ∼ 0) indicates that no single variable plays a substantial role in mediating infection dynamics [47]. Using the *Anova* function in the R package car [48], we assessed significance of the effects using Wald χ□2 statistics for the winning model.

**Supplementary Table 1**. Model selection for prevalence of *M. ozzardi* across demographic variables. Variables include age, sex, education, ethnicity, and location. Global = focal model, m8 = winning model.

### Protocol and data availability

All pipelines for statistical analysis and data visualization are available at https://github.com/zamanianlab/Mansonella-ms.

Raw read data used for assembly of mitogenomes of *M. ozzardi* isolates Moz-Col-195, Moz-Col-204, Moz-Col-220 and Moz-Col-239 are submitted under NCBI BioProject XYZ. The accession numbers of the assembled mitogenomes are XYZ, XYZ, XYZ, XYZ respectively. The mitogenomes of isolates Moz-Brazil-1 and Moz-Venz-1 were assembled from previously reported raw read datasets [16] available from NCBI BioProject PRJNA917722 and PRJNA917766 respectively. The corresponding GenBank accessions for these mitogenomes are XYZ for Moz-Brazil-1 and XYZ for Moz-Venz-1.

## Results

### Parasite Prevalence

To assess *Mansonella* prevalence within our study population, we deployed a species-specific loop-mediated isothermal amplification (LAMP) assay [30] and compared diagnostic results with microscopic examination of thin blood smears. Across the first survey (samples 1-117), blood LAMP results show a higher prevalence of *M. ozzardi* infections (54/115, 46.9%) than single thin smear microscopy (16/104, 15.3%) and LAMP carried out using DNA from matched plasma (31/115, 26.9%) (**Figure 2 A**). Consensus blood LAMP results were derived from experiments involving DNA extractions from independent aliquots of blood as well as replicates of LAMP assays carried out in two laboratories using DNA from a single extraction (**Supplementary Figure 1 A**).

**Figure 2.**
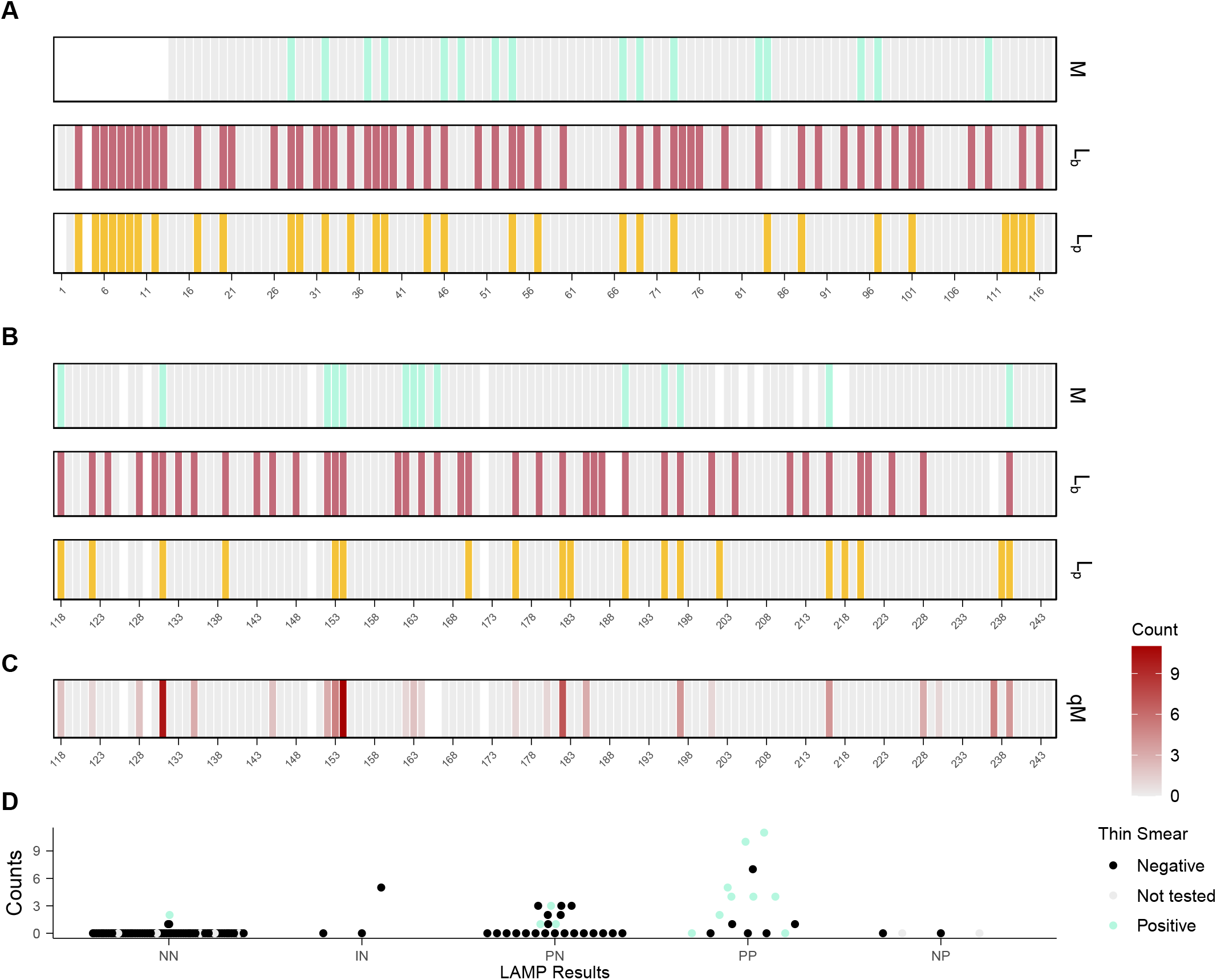
**A)** Diagnostic results derived from survey one blood samples (1-117) for in-field microscopy smears (M), LAMP using DNA extracted from blood (L_b_), and LAMP using matched DNA extracted from plasma (L_p_). **B)** Diagnostic results derived from surveys two and three (samples 118-244) for in-field microscopy smears (M), LAMP using DNA extracted from blood (L_b_), and LAMP using matched DNA extracted from plasma (L_p_). **C)** Quantitative microscopy counts (qM) derived from surveys two and three blood samples (118-244). **D)** Quantitative microscopy counts of microfilariae stratified by L_b_ and L_p_ results and coloured by microscopy smear results. (NN = negative L_b_ and negative L_p_, IN= inconclusive L_b_ and negative L_p_, PN = positive L_b_ and negative L_p_, PP = positive L_b_ and positive L_p_, NP = negative L_b_ and positive L_p_). Grey = negative for *M. ozzardi*; White = no result (A-C). LAMP assay data are specific to *M. ozzardi*.

Treating the blood LAMP as the truth case, thin smear microscopy shows a 31% sensitivity and plasma LAMP shows a 50% sensitivity. For the 30 samples where blood LAMP was positive and microscopy was negative, lower time (Tt) which is indicative of more parasite material being present in semi-quantitative LAMP (sq-LAMP) were observed (**Supplementary Figure 1 B-C**). We therefore hypothesized that the discrepancy between LAMP results and microscopy is likely associated with lower titers of microfilariae in venous blood leading to thin smear false negatives [49] or occult (amicrofilaremic) infections [21] that can only be detected using molecular methods. In both cases, we would expect the blood LAMP assay to display better sensitivity.

Subsequent surveys (samples 118-186; samples 187-244) were used to probe potential associations between microfilaremia and *Mansonella* diagnostic results by incorporating quantitative microscopy. Blood LAMP assays showed an *M. ozzardi* prevalence of 33% (40/120). Among the 14 samples which tested positive in microscopy, 13 also tested positive in LAMP. Plasma LAMP (16%) and thin smear microscopy detected a lower prevalence (12%), reflecting the pattern observed in the first survey (**Figure 2 B**). Quantitative microscopy detected mf in 23/122 samples (19%), ranging from 0-11 mf/blood spot (**Figure 2 C**). Samples with the highest mf titers were most likely to be positive across blood and plasma LAMP assays and thin smears, while samples with lower mf titers were more likely to be negative in blood smears and only detected in the blood LAMP assay (**Figure 2 D**). This lends support to the hypothesis that low mf titers is a driver of false negatives in both thin smears and plasma LAMP assays.

To explore whether occult infections could also be a factor contributing to the positive blood LAMP samples which had negative microscopy results, we re-sampled blood from six individuals that fit this pattern. A larger volume of blood (1 mL) was filtered to identify mf that could be circulating at titers not routinely detectable via thick and thin smear microscopy. All six samples were still microscopy negative, supporting the likelihood of occult infections. Whole blood LAMP was the most sensitive diagnostic test and identified a prevalence of 40% for *M. ozzardi* across all three surveys (**Table 1**).

**Table 1.** Summary and comparison of *M. ozzardi* diagnostic results for the entire study population.

|  | Microscopy Smear /<br>LAMP-Plasma Positive | Microscopy Smear /<br>LAMP-Plasma Negative | No Result | Total |
| --- | --- | --- | --- | --- |
| LAMP-Blood<br>Positive | 27 / 42 | 57 / 52 | 10 / 0 | 94 |
| LAMP-Blood<br>Negative | 3 / 8 | 129 / 132 | 9 / 1 | 141 |
|  | 30 / 50 | 186 / 184 | 19 / 1 | 235 |

**Supplementary Figure 1 A)** Diagnostic results derived from survey one blood samples (1-117) for four replicates of LAMP using DNA extracted from blood samples (L_b_) across two laboratories. Grey = negative for *M. ozzardi*. White = no result. **B)** Diagnostic results derived from survey one blood samples (1-117) for semi-quantitative LAMP (Tt). ND = not detected **C)** Semi-quantitative LAMP Tt counts (minutes) stratified by L_b_ and L_p_ results and colored by thin smear results. Dashed lines indicate Tt cuttoffs (> 1, <30 minutes = positive sq_LAMP). (NN = negative L_b_ and negative L_p_, PN = positive L_b_ and negative L_p_, PP = positive L_b_ and positive L_p_, NP = negative L_b_ and positive L_p_). LAMP assay data are specific to *M. ozzardi*.

### Population Demographics

The three surveys consisted of individuals who reside in thirteen different communities situated between -02°50’14.9496” north latitude and -03°52’39.1980” south latitude; and - 069°44’10.7880” and -070°35’52.2240” west longitude (Figure3 A). A majority of our sampled population were centered in Puerto Nariño (n = 136) (Table 2), where the prevalence of *M. ozzardi* as determined by blood LAMP is 44%. The next largest sampled communities in our survey, 12 de Octubre (24%; n = 25) and San Pedro de Tipisca (14%; n = 21), had lower prevalence. There was a higher prevalence of *M. ozzardi* mf identified via blood LAMP in males (49%; n = 84; mean age = 40.1 years) compared to females (32%; n = 148; mean age = 34.7 years). The mean age of *M. ozzardi* positive individuals was higher than the mean age of negative individuals in females (Wilcox: p = 0.026) but not males (Wilcox: p = 0.07) (Figure 3 B). To further examine differences between infection prevalence across demographic variables, we assessed significance of the fixed effects using Wald χ□2 statistics and identified education and place of residence as drivers of prevalence (education binomial GLM: χ□2 = 16.2647, p = 0.01240, case origin GLM: χ□2 = 13.6850, p = 0.03336). However, given the limitation of the data we were unable to confirm significant differences within these groups due to unequal and small sample sizes amongst subsets.

**Table 2.** Summary of Demographic and diagnostic data. Data includes a subset of the total study population where all summarized demographic information was provided on the questionnaire. (Age in years).

|  | Young Adult<br>18 - 32 | Adult<br>33 - 54 | Mature Adult<br>55 - 89 | Total |  |  |  |  |
| --- | --- | --- | --- | --- | --- | --- | --- | --- |
| Age | 101 | 104 | 29 | 234 |  |  |  |  |
| Sex (Female / Male) | 69 / 32 | 68 / 36 | 12 / 17 | 149 / 85 |  |  |  |  |
| *M. ozzardi* (L<sub>b</sub> / L<sub>p</sub> / Microscopy) | 31 / 16 / 9 | 42 / 24 / 16 | 20 / 10 / 5 | 93 / 50 / 30 |  |  |  |  |
|  | Primary | High School | Technical | University | Other | Not Reported |  |  |
| Education | 59 | 81 | 52 | 16 | 2 | 24 |  |  |
|  | Bora | Cocama | Senu | Ticuna | Tokami | Multiethnic | Not Reported |  |
| Ethnicity | 1 | 5 | 1 | 151 | 1 | 3 | 72 |  |
|  | Lomas Lindas | 12 De Octubre | San Francisco | Villa Andrea | Puerto Narino | San Pedro De Tipisca | Other | Not Reported |
| Location | 11 | 25 | 9 | 15 | 136 | 21 | 14 | 3 |

**Figure 3.**
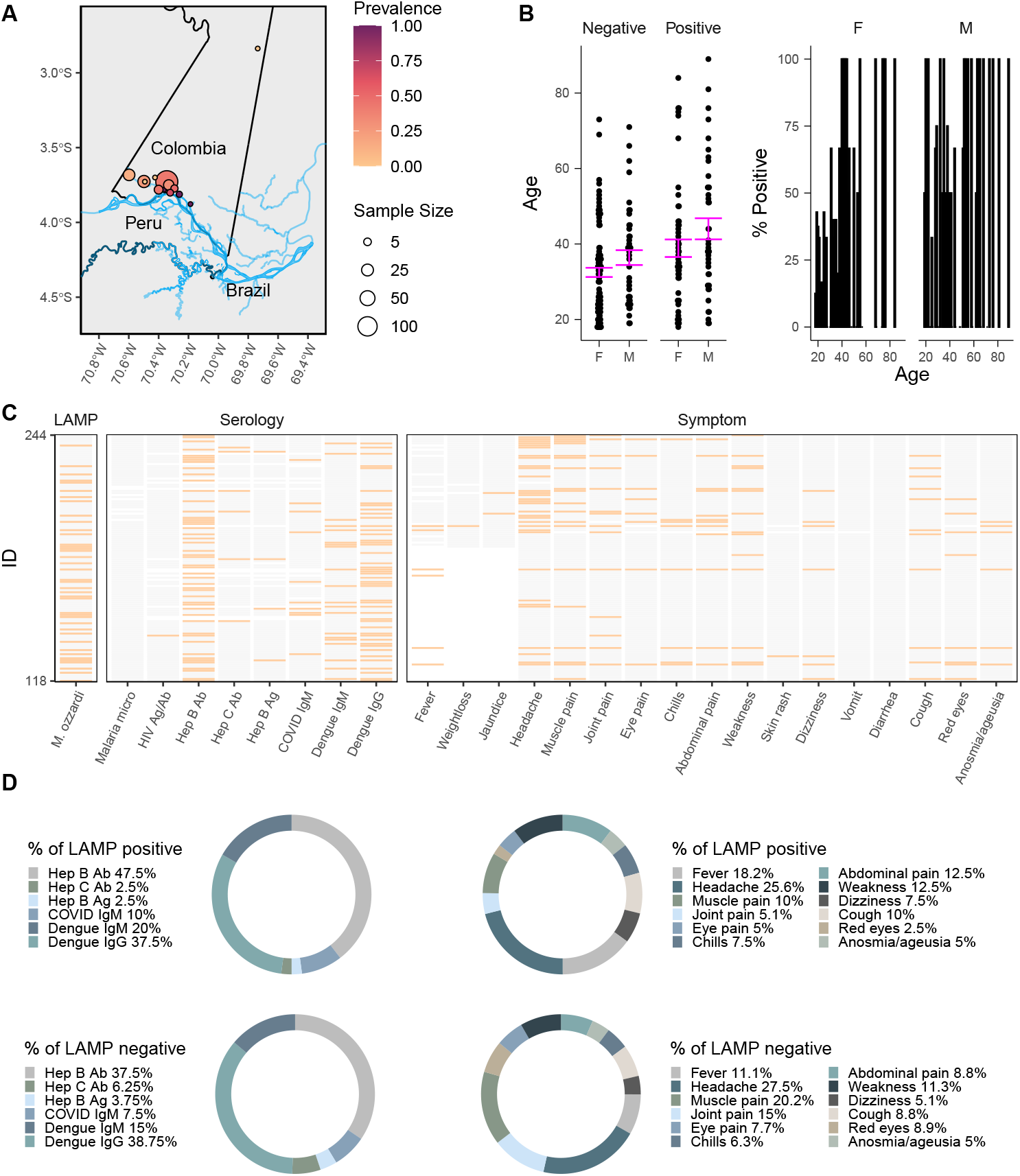
**A)** A map of the study region in the Amazonas Department of Colombia depicting the community where individual participants reside. Point size reflects the number of samples collected and color represents the prevalence of *M. ozzardi* infection for each community. **B**) *M. ozzardi* prevalence as determined by blood LAMP assay stratified by sex and age (95% CI) (left) and histogram of *M. ozzardi* prevalence distribution by age (right). F = female, M = male **C**) *M. ozzardi* infection status and reported symptoms and serological test results for survey two and three individuals (n = 120). Orange = positive, present; Grey = negative, not-present; white indicates missing data. **D**) Donut plots reflecting relative fractions of *M. ozzardi* L_b_ positive and L_b_ negative individuals who self-reported symptoms and lab confirmed serological test results.

Clinical histories were collected and additional serological tests were carried out for survey two and three samples (n = 117) (**Figure 3 C**) and compared across *M. ozzardi* blood LAMP positive and negative individuals (**Figure 3 D**). Symptoms and serology results that were differentially reported (abdominal pain, fever, joint pain, muscle pain, Hepatitis B antibodies, and Hepatitis C antibodies) between blood LAMP positive and negative individuals were examined further to determine if fixed effects or *M. ozzardi* infections were more likely to explain these data. We assessed significance of the fixed effects and blood LAMP results using Wald χ□2 statistics and determined that *M. ozzardi* infection status did not significantly correlate to reported symptoms or serology results. Fixed effects including sex and age were identified as potential drivers of some of the reported symptoms and serology results but unequal and small sample sizes amongst subsets of data limited our post-hoc analysis.

### Genomic Analysis

Whole genome sequencing was performed on 4 samples (ID numbers 195, 204, 220 and 239) which tested positive in the *M. ozzardi* specific blood LAMP assay. Only sample 239 had detectable microfilariae in the quantitative microscopy assay. While a mapping analysis of the sequencing data showed that over 94% of the reads were from the human host, 1 to 6 % of reads mapped to the reference *M. ozzardi* genome assembly and to the *Wolbachia w*Moz genome assembly, although at a low coverage (**Supplementary Figure 2**).

Sufficient read coverage was obtained for successful assembly of complete, circular mitogenomes for each isolate. The assembled mitogenomes displayed more than 99.6% sequence identity to each other and to previously reported *M. ozzardi* mitogenomes from Brazil [21,33], as well as to the newly assembled mitogenomes from isolates Moz-Brazil-1 and Moz-Venz-1 obtained from Brazil and Venezuela respectively [16]. A phylogenetic analysis of all *M. ozzardi* mitogenomes in conjunction with *M. perstans* mitogenomes from Brazil [21] and Cameroon [38] showed a distinct *M. ozzardi* clade with very short branch lengths between various isolates and a clear separation from the *M. perstans* clade (**Figure 4**). Together, these results confirm the presence of *M. ozzardi* infection in these individuals.

**Figure 4.**
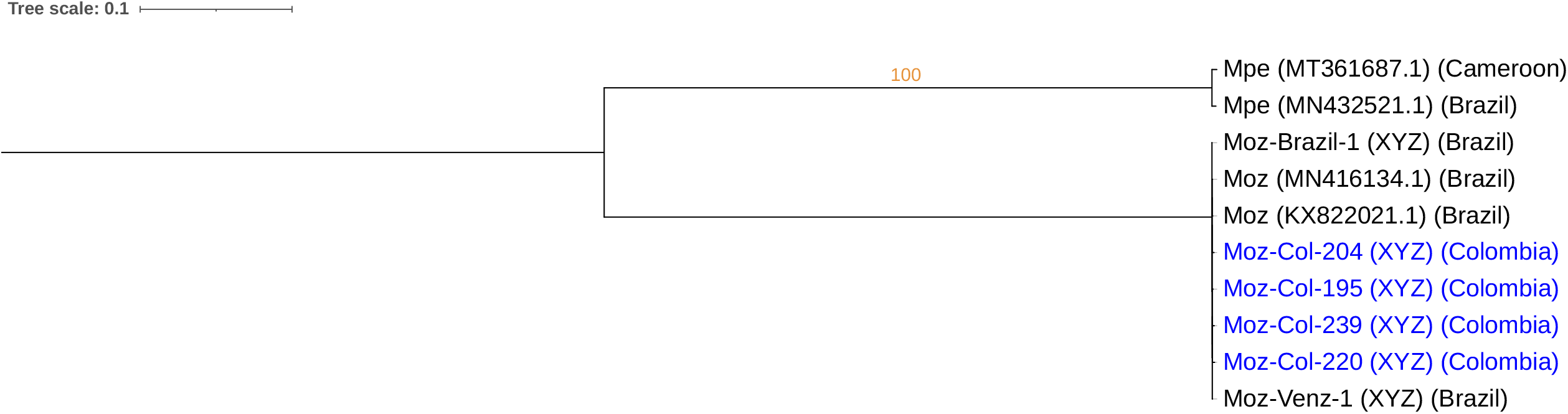
Maximum-likelihood tree based on whole-mitogenome alignments of various *M. ozzardi* and *M. perstans* isolates. Abbreviation Moz and Mpe and indicate *M. ozzardi* and *M. perstans* respectively. The GenBank accessions are indicated in parenthesis next to isolate names, followed by the country of origin in the second parenthesis. Blue colors mark the isolates sequenced in this study. The DNA substitution model HKY+F+I was found to be the best fit according to Bayesian Information Criteria in ModelFinder. Values of ultrafast bootstrap support calculated with 1000 replicates is shown for branches with value higher than 80.

**Supplementary Figure 2**

**A)** Sequencing read coverage from different *M. ozzardi* isolates on the reference nuclear contigs from Moz-Venz-1 genome assembly. **B)** Sequencing read coverage from different *M. ozzardi* isolates on the *Wolbachia w*Moz reference genome assembly.

### Cryopreservation of Blood-Dwelling Parasite Stage

As a first step towards establishing the lifecycle of *M. ozzardi* in a laboratory environment, we sought to functionally cryopreserve microfilariae and observe viable and active worms after thawing samples. We collected peripheral blood from individuals who were positive for *M. ozzardi* via blood LAMP assay and thin smear microscopy in our initial survey. Blood samples were mixed with dimethyl sulfoxide (DMSO) and kept at -80°C. We observed revitalization of microfilariae upon thawing at 37°C, whereby motility increased over the first 24 hour incubation period before steadily declining (**Figure 5 A**). Videos showed viable and healthy microfilariae (**Figure 5 B**) that may allow for membrane feeding of lab colonies of *Culicoides* and the production of infective-stage larvae (L3) in a laboratory setting.

**Figure 5.**
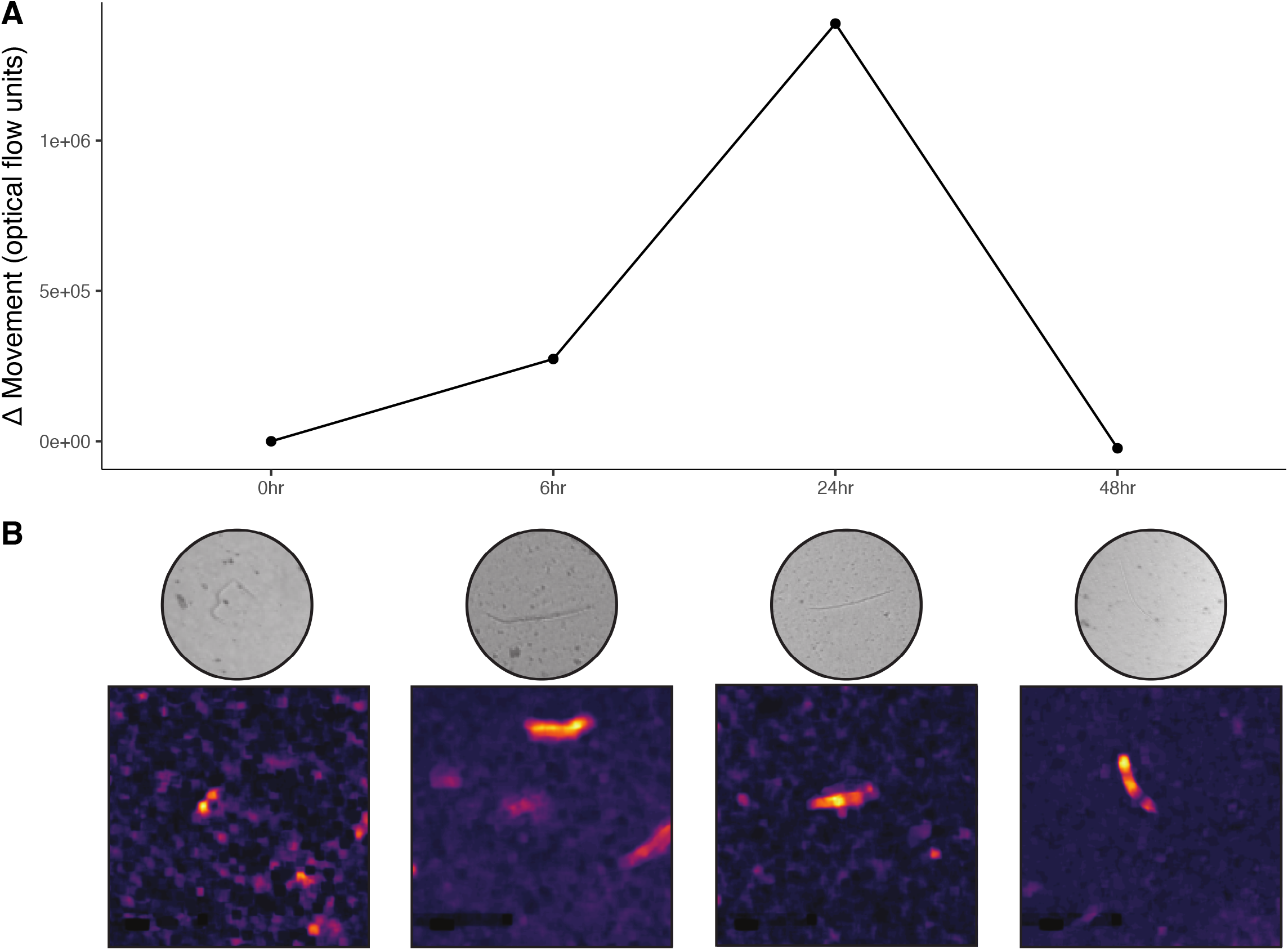
A) Motility of cryo-preserved *M. ozzardi* microfilariae over 48 hours as measured by an optical flow algorithm. **B)** Brightfield images and optical flowmaps of videos used for motility analysis of thawed cryopreserved *Mansonella* samples aligned with the timepoints in (A). Brighter regions of the flowmaps reflect areas where more parasite movement was detected.

## Discussion

We conducted three seroprevalence surveys to better map the distribution of *Mansonella* infections among indigenous communities in the Amazon basin of Colombia. Blood smear microscopy identified microfilariae in 12.7% of sampled individuals, which falls within the range of historical studies in the region [23,24,50]. Blood LAMP assays detected *M. ozzardi* DNA in 40% of sampled individuals, in closer agreement with more recent PCR-based surveys in the Amazon region of neighboring countries [26,28,51,52]. Our survey data adds to the growing body of evidence that microscopy-based approaches can drastically underestimate the true prevalence of global mansonellosis [5,14,26–30,51–53].

Molecular diagnostic assays can be leveraged to capture a broader range of infection states, including low microfilaremias or afilaremic (occult) infections [21]. The recovery of complete mitogenomes confirms the presence of *M. ozzardi* DNA in blood samples that previous microscopy diagnostics could not detect, providing further confidence in LAMP diagnostic results. The distribution of *M. ozzardi* adults within the human body is not fully resolved [54], but it is presumed that they commonly take residence in subcutaneous tissues and possibly the serous cavities. These larger stages are the likely source of blood-detectable nucleic acids in molecular diagnostic assays. LAMP assays provide a sensitive, specific, and potentially cost-effective approach for the surveillance of filarial nematode infections in human and vector populations [30,55–57]. The development of direct LAMP assays [58] not reliant on DNA extraction will add to the convenience of this approach in endemic settings.

The establishment of a cryopreservation protocol can facilitate the eventual rearing of *M. ozzardi* in laboratory settings, including controlled blood-feeding of reanimated microfilariae to susceptible colonies of biting midges. In vitro phenotyping of *Mansonella* drug responses [59] can help resolve the genetic and molecular basis for observed differences in antifilarial drug susceptibility across species [9,10,12,13,15,60] and open avenues for the screening and discovery of new therapeutic leads for mansonellosis.

*Mansonella* is neglected even among neglected pathogens due to a lack of investment in studying its biology and potential health impacts [4,8,61]. More complete demographic and clinical data are needed to determine *M. ozzardi* pathogenicity and assess risk factors. Adult worms disperse somewhat randomly into various tissues and body cavities, the consequences of which can vary considerably across individuals but have historically been summarized as relatively non-pathogenic. Although there are growing clinical case reports suggesting appreciable pathogenicity [4], it is difficult to streamline causal associations with more subtle health impacts. The current established threshold for addressing human filariasis is a causal association with significant morbidity, including blindness and physical disfigurement. Investigation of subclinical or secondary impacts of infection would shift the threshold of potential human clinical concern in mansonellosis-endemic regions to the same threshold that often triggers action as it relates to subclinical nematode infections of livestock and companion animals. We expect that improved diagnostic tools, growing genomic resources, and methods to study *Mansonella* in laboratory settings will facilitate this goal.

## Data Availability

All diagnostic data produced in the present work are contained in the manuscript in de-identified fashion. Genomic data were filtered for reads that map to pathogens of interest and are deposited in the NCBI SRA.

https://github.com/zamanianlab/Mansonella-ms

## Acknowledgements

The authors thank the communities, the Secretaria de Salud Departamental del Amazonas, Alcaldia de Puerto Nariño, E.S.E Hospital San Rafael de Puerto Nariño, and the Laboratorio Departamental de Salud Pública de Leticia for their contributions to this study. We sincerely thank Julian Rodriguez and Corporacion Corpotropica for their assistance and coordination of activities at the study sites. In addition, we would like to thank the staff of One Health lab and auxiliary personnel at all the study sites for their assistance. Thanks to the Abbott Pandemic Defense Coalition for the provision of laboratory testing kits.

