## Supplementary figures and images for "Molecular surveillance detects high prevalence of the neglected parasite *Mansonella ozzardi* in the Colombian Amazon"

### Supplementary Figure 1

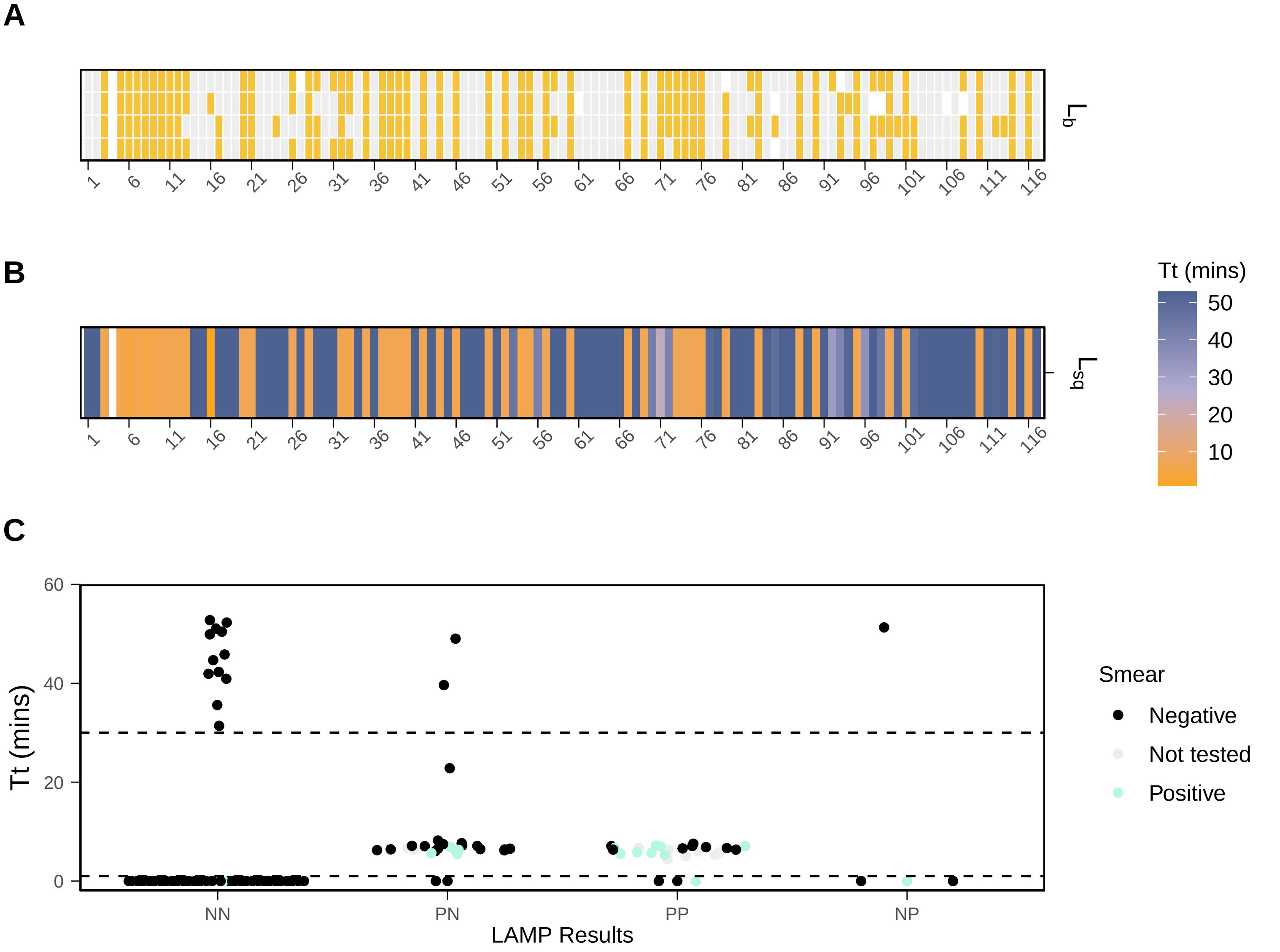

### Supplementary Figure 2

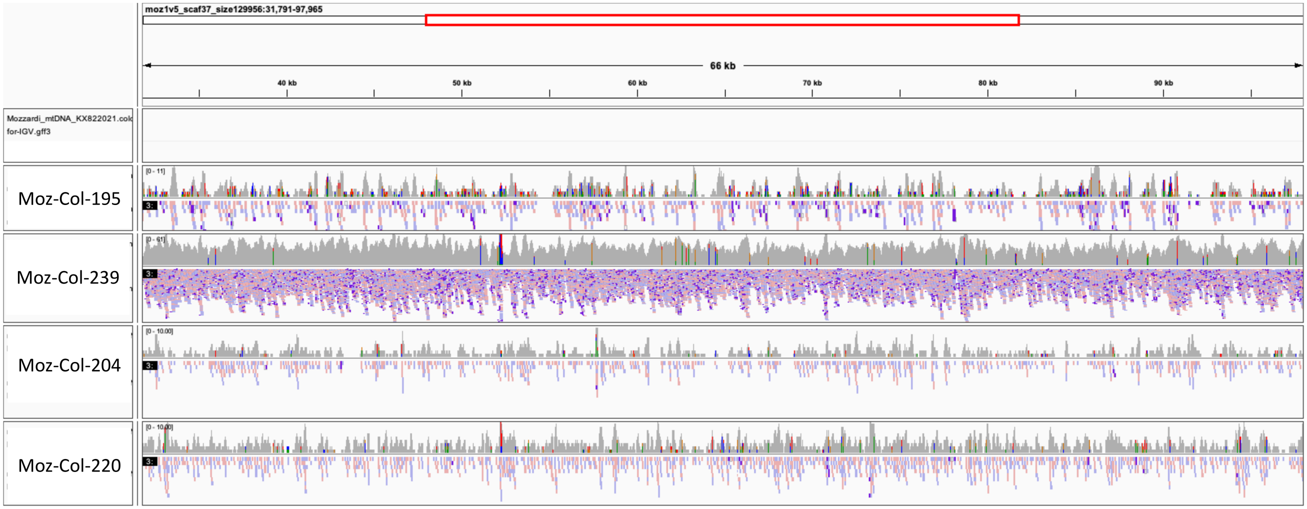

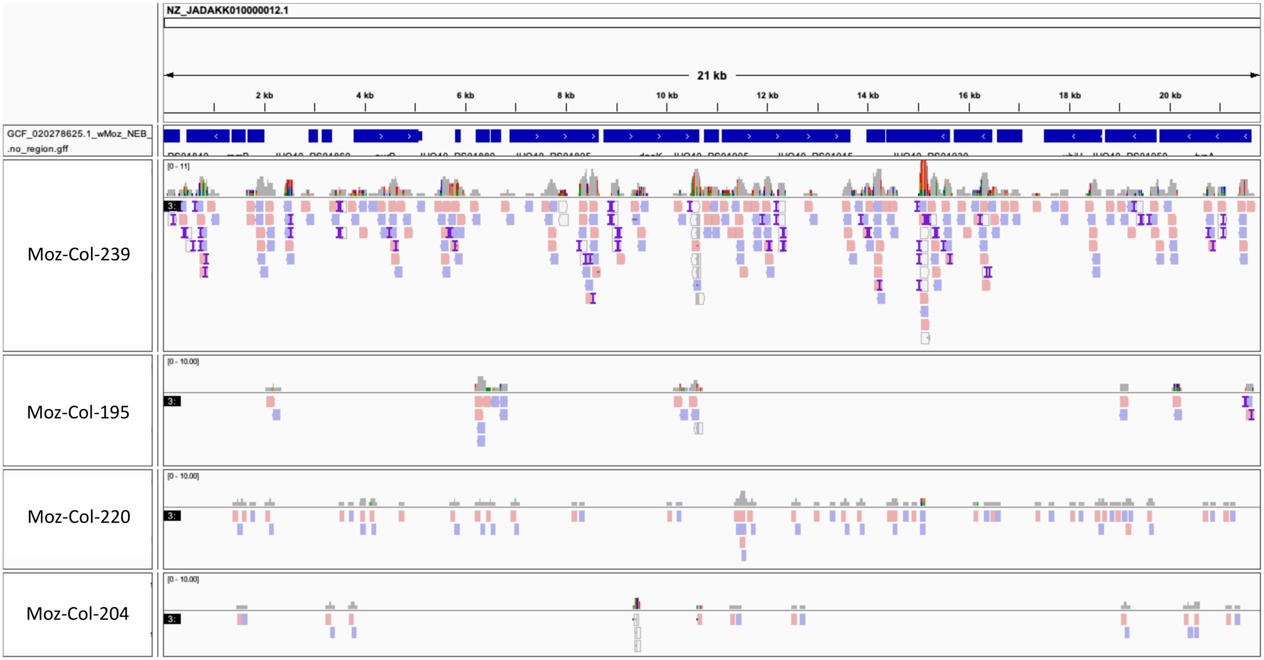
