## Supplementary Table1 for "Molecular surveillance detects high prevalence of the neglected parasite *Mansonella ozzardi* in the Colombian Amazon"

Table 1: Model selection table on Prevalance of infection

| Model | K | AIC | Delta AIC | AIC weight | log-Likelihood |
|---|---|---|---|---|---|
| m8 | 14 | 145.41 | 0.00 | 0.54 | -58.71 |
| m7 | 15 | 147.60 | 2.19 | 0.18 | -58.80 |
| m2 | 16 | 149.02 | 3.61 | 0.09 | -58.51 |
| m11 | 18 | 149.43 | 4.01 | 0.07 | -56.71 |
| m5 | 19 | 150.33 | 4.92 | 0.05 | -56.17 |
| m9 | 13 | 151.55 | 6.13 | 0.02 | -62.77 |
| m6 | 10 | 151.72 | 6.30 | 0.02 | -65.86 |
| m4 | 20 | 153.38 | 7.97 | 0.01 | -56.69 |
| m10 | 14 | 154.25 | 8.83 | 0.01 | -63.12 |
| m1 | 21 | 154.30 | 8.89 | 0.01 | -56.15 |
| m3 | 15 | 154.49 | 9.08 | 0.01 | -62.25 |
| global | 23 | 155.58 | 10.17 | 0.00 | -54.79 |
